# A Comparative Study on the knowledge and attitude of COVID-19 among Urban and Rural populations of Bangladesh

**DOI:** 10.1101/2021.08.10.21261843

**Authors:** Md. Kamal Hossain Ripon, Noor Muhammad Khan, A E M Adnan Khan, Rana Ahmed, Safia Afrin, Md. Abu Sayed, Md. Mizanur Rahman Moghal

## Abstract

**Aim:** This study is aimed to identify the awareness and behavioral perspective on COVID-19 between urban and rural people of Bangladesh during the period of outbreak.

**Methods:** A community-based cross-sectional study was conducted in 30 districts of Bangladesh, where 322 participants from urban and 312 from rural area. Participants were selected through convenience sampling.

**Results:** Rural people are found to be much more inter connected to receive information from neighbor. Regarding the incubation periods and the general symptoms, knowledge differs significantly from urban to rural. Even their precautionary and transmission knowledge is found to associate in most of the cases. During this outbreak, urban people significantly increase their religious habits and also believe that there will some major change of life after outbreak.

**Discussion:** The study reflected that health education program needed to aware about COVID-19 in both urban and rural in Bangladesh that helps in formulating and executing communication and outbreak management.

## Introduction

In 1960, corona virus was first appeared and until 2002, the world considered it as a nonfatal and relatively simple virus. The outbreak of 2002-2003 in China later spread many other countries including United States of America with high mortality rates. After massive fatality, Centers for Disease Control and Prevention and World Health Organization (WHO) declared a state of emergency in 2004 [1-3].An unknown case of pneumonia was reported which clinical symptoms were similar to usual viral pneumonia in Hubei province, China, in December 2019 [4]. The pneumonia was named by the World Health Organization (WHO) and the International Committee on Taxonomy of Viruses as “COVID-19” and ‘Severe Acute Respiratory Syndrome Coronavirus2’ (SARS-CoV-2) respectively [5]. It is now a pandemic and an international emergency of public health for all over the countries, should step forward to prevent COVID-19 spread called by World Health Organization (WHO) on January 30 [6, 7].

The COVID-19 was confirmed to spread in Bangladesh on March 2020. The first three known cases were reported by the country’s Institute of Epidemiology, Disease Control and Research (IEDCR) on 8 March 2020 [8]. Within 4 May, there are a total of 10143 confirmed cases, 182 deaths and Case Fatality Rate (1.79%) in the country [9].

Bangladesh first imposed nationwide lockdown from March 26 and extended several times for the consequence the Ministry of Public Administration again issued a notification on 4 May, 2020 to extend the general holiday and close all schools, colleges and universities until 14 May, followed by a weekend 15-16 May except all emergency services to resist the spreading of COVID-19 [10].Government of Bangladesh bound to withdraw lockdown due to the economic distress related to suicidal incidences around that time [11, 12].

Within the last two weeks of march, 2021, the number of infection and death is tremendously increases and high-risk zone gave a hyper jump from 10 to 38 which is more than half country’s 64 districts, according to IEDCR data [13]. From June, 2021, both the number of infection and death dramatically increases and government reimposed a strict lockdown nationwide from July 1-13 and ease the lockdown from July 15-22 for the biggest festival Eid al-Adha and again resume the strict lockdown from July 23 to next two weeks [14-16].

Overall, 1,994,752 infected cases and 19,779 deaths reported in Bangladesh [17] and globally more than 196,002,202 people was infected and 4,193,301 confirm death by COVID 19 on 28th July, 2021 [18].During the period of outbreaks, general people need instant information, a group of population is experience fear, discrimination and stigmatization required special care [19, 20].Furthermore, after the outbreak of severe acute respiratory syndrome (SARS), Middle East respiratory syndrome (MERS), and Ebola, it was recommended that the knowledge and attitudes is connected with the intensity of panic emotion regarding the infectious diseases which make further difficulties to prevent the spread of the diseases [21-26]. While the illness and death are significant, general public or specific communities suffer from fear which make them delay asking help and remain undetected that is very hazardous for controlling transmission during the outbreak of infectious diseases [25]. After the outbreak, the prevalence of post-traumatic stress disorder (PTSD) and major depression of general people increased up to 41% and 7% respectively [27].

At this critical situation, it is vital need to understand the public’s awareness of COVID-19 in Bangladesh to facilitate the management of outbreak. In this study, we investigate the knowledge and attitude towards COVID-19 of both urban and rural residents of Bangladesh during this rise period of outbreak to provide the legislators actual field-based data and to support them in the management of this pandemic.

## Methods

To capture the attitude toward COVID-19 among the people of Bangladesh, a community-based cross-sectional study was conducted over a short period (March 2020 to April 2020) during the rise period of outbreak of COVID-19. We have collected the data from 30 convenient districts out of 64 districts. Due to lockdown situation in Bangladesh, it was very hard to collect the data from all the districts. Total 634 participants are encountered in the survey, where we tried to make equal representation of urban and rural people. Respondents were also selected from each district based on their availability to us. In a ward, we have used convenience sampling, a non-probability sampling technique, in selecting the respondents from the people of Bangladesh. This sampling technique is also known as accidental sampling in many literatures. Convenience sampling involves the sample being drawn from that part of the population that is close to hand. Though it increases the selection bias, it was the only efficient way of collecting data from the people of Bangladesh in lockdown days.

The questionnaire was developed based on the knowledge about COVID-19. We designed the questionnaire into several sections including Transmission, Sign & Symptoms, Precautions, Treatment, Mental Health, and socio-demographic status of the respondents. Each part is a mirror image of the knowledge about COVID-19 except first and the last part which contains the demographic characteristics and mental health of the participants respectively. In the second section, the questions were designed to reflect the basic knowledge about COVID-19 among rural and urban populations. Knowledge about the transmission is most important part to resist the COVID-19, which was measured through the questions in section three. Similarly, questions in section four reflect the knowledge regarding sign and symptoms among the mentioned populations. Another most important part in the knowledge of precaution was measured through the questions in the fifth section and a prime factor of contamination rate among them. Questions in the sixth section were designed regarding the treatment option against COVID-19. Finally, our study observed the mental health of the participants during and after the pandemic through the questions of the last section.

One of the uses of the *χ*^2^ statistic is in contingency (dependence) testing where n randomly selected items are classified according to two different criteria, such as when data are classified on the basis of two factors (row factor and column factor), where the row factor has r levels and the column factor has levels. We use chi square test (*χ*^2^)in our research to test the hypothesis that the two or more factors are independent. P values of the chi square test are presented in the table.

## Results

Table 1 summarizes the participants according to their demographic characteristics. There is almost equal representation of urban and rural people in our data set, where female participants are found to slightly more convenient than male. We have collected the data from those people who have age 13 or above. Among them, 94% are found to be educated as they have completed their secondary school certificate. Majority of our respondents are from the middle-class family or higher which ensures that they have the sufficient resources to gather knowledge about COVID-19. Unfortunately, we didn’t find any survivor respondents.

**Table 1:**
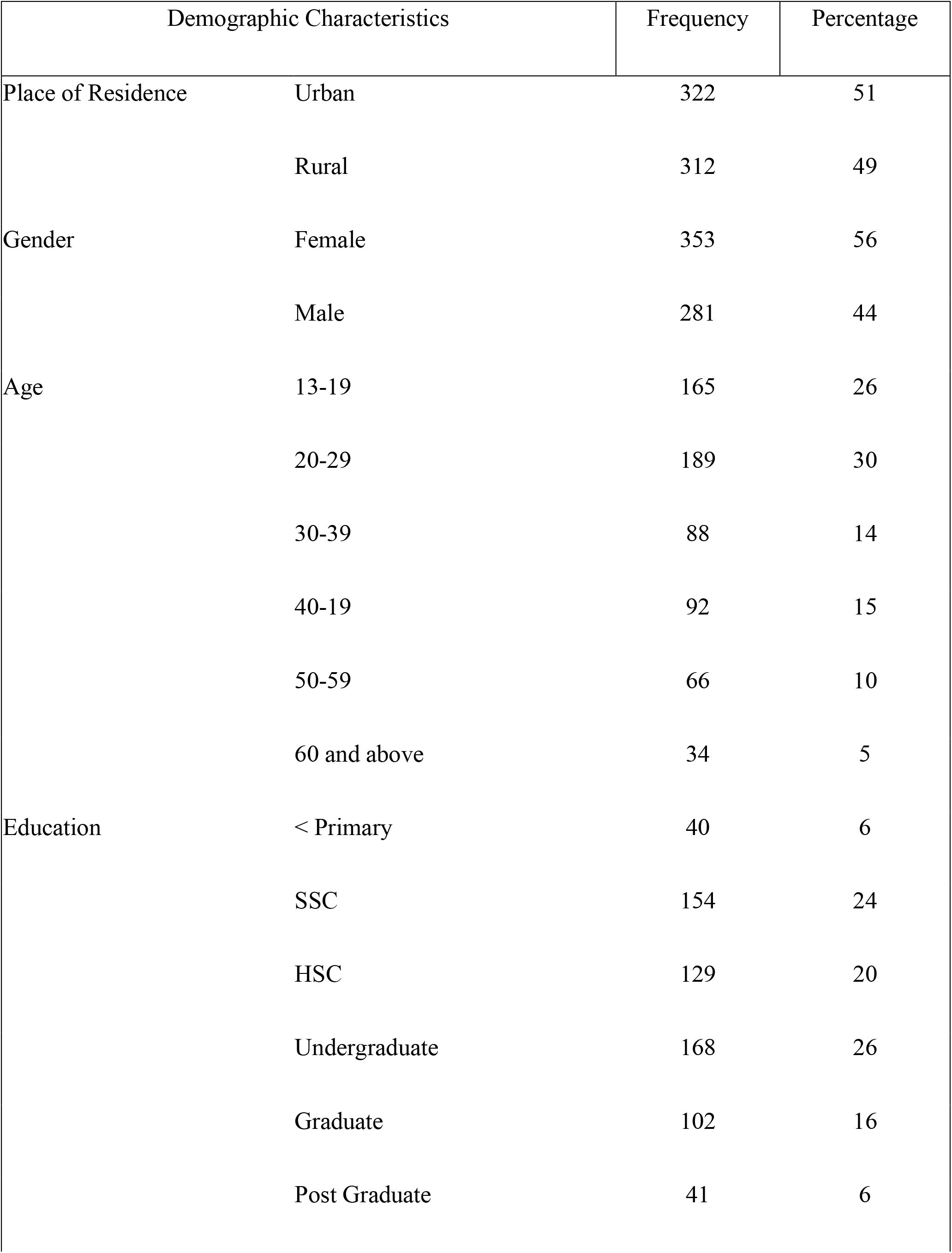
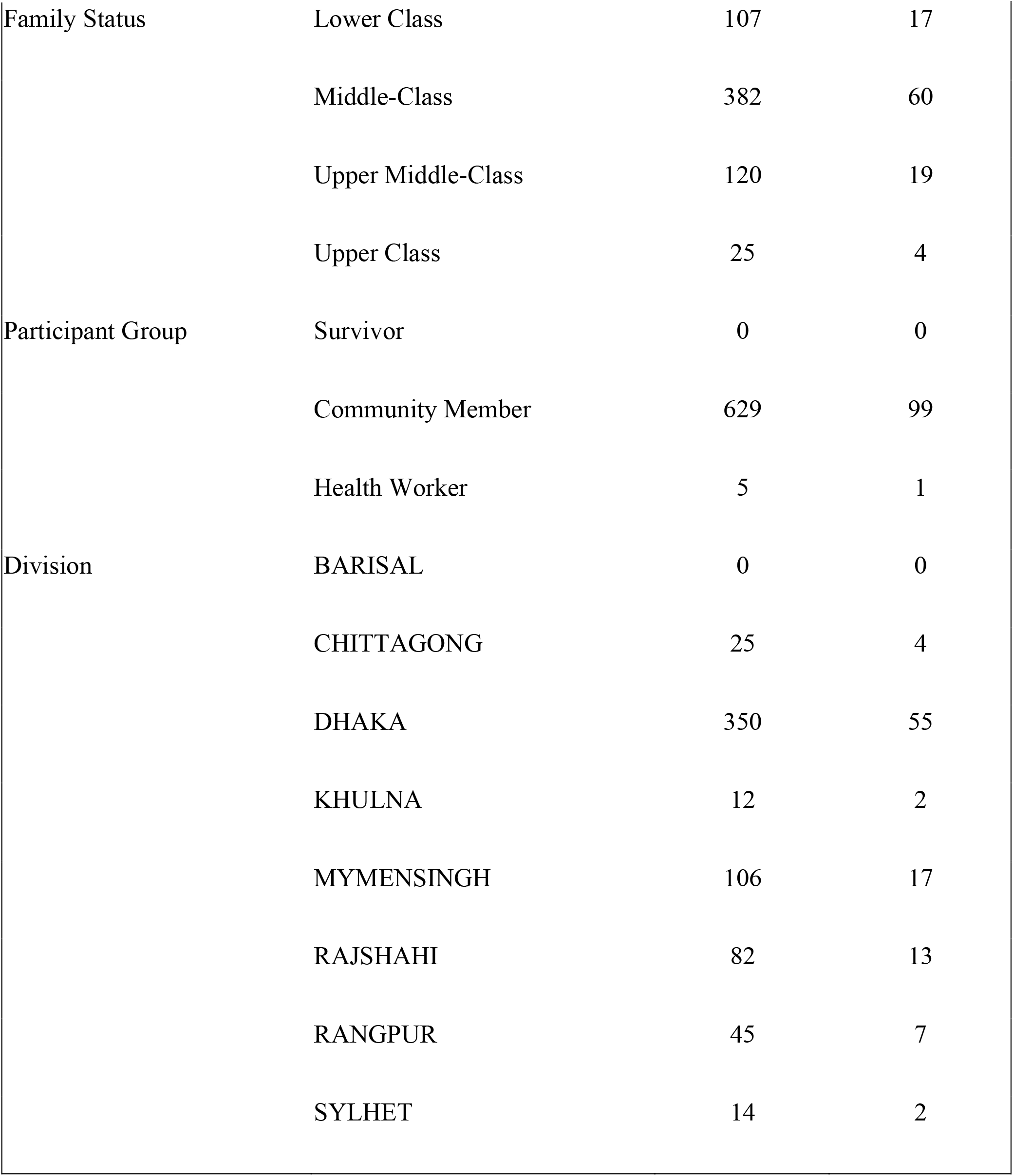
Population according to Demographic Characteristics

Figure 1 represents the distribution of urban and rural people who have heard about the corona virus, have known about what is corona virus and finally whether they are familiar with the causes of COVID-19 before attending the survey study or not.

**Figure 1:**
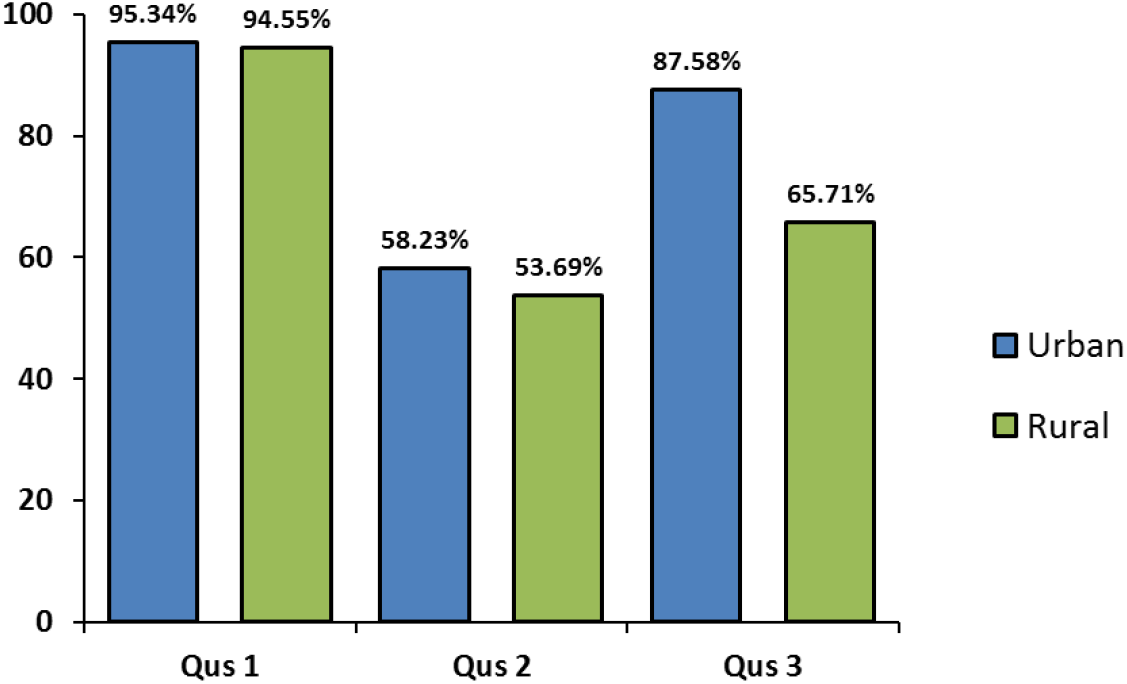
Distribution of urban and rural people according their general knowledge about COVID-19.

People in urban and rural bear similar general information about COVID-19 on their mind except the cases, where urban people are more knowledgeable about the causes of this novel virus than the rural people.

Figure 2 represents that how the participants from the urban and rural, known about the information of COVID-19. Mass media is key source of knowledge about COVID-19 in both urban and rural area of Bangladesh, where Friends are the secondary source of information there. Compared to urban, people in rural get much information about this virus from their neighbors.

**Figure 2:**
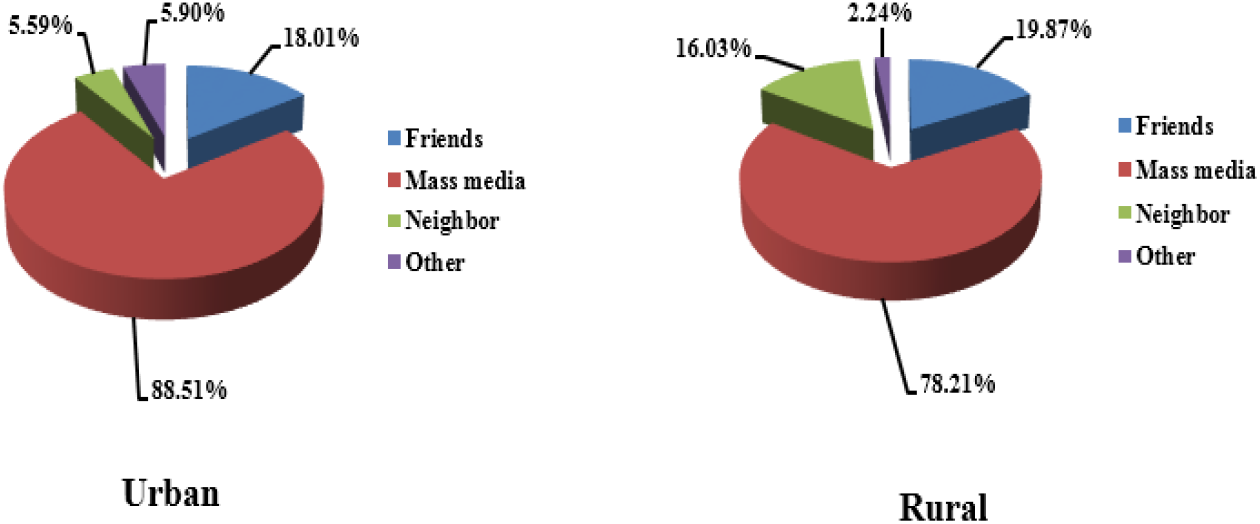
Distribution of knowledge source about COVID-19 in urban and rural.

Table 2 reveals that there no statistically significant difference on general knowledge about COVID-19 between urban and rural people in Bangladesh.

**Table 2:**
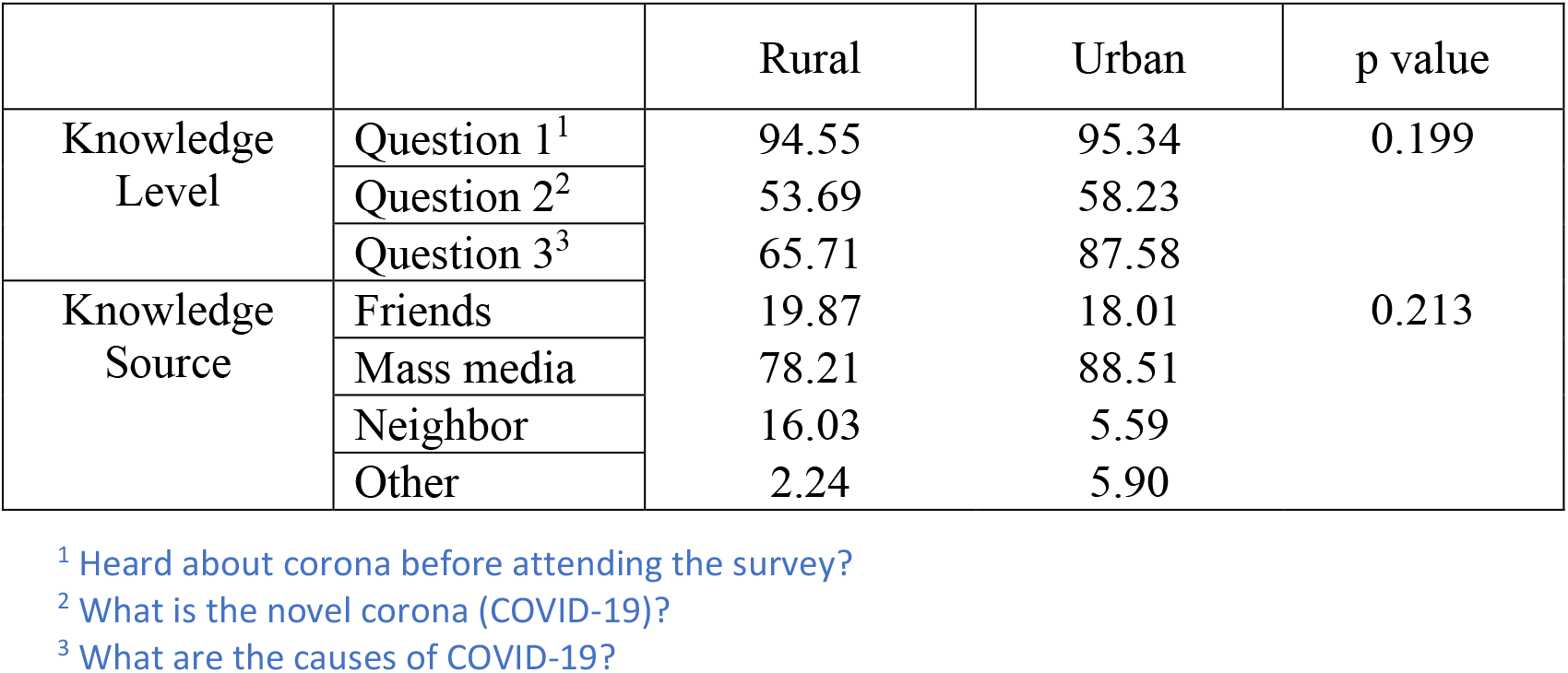
Distribution of knowledge level and its source about COVID-19 in urban and rural.

|  |  | Rural | Urban | p value |
| --- | --- | --- | --- | --- |
| Knowledge Level | Question 1<sup>1</sup> | 94.55 | 95.34 | 0.199 |
|  | Question 2<sup>2</sup> | 53.69 | 58.23 |  |
|  | Question 3<sup>3</sup> | 65.71 | 87.58 |  |
| Knowledge Source | Friends | 19.87 | 18.01 | 0.213 |
|  | Mass media | 78.21 | 88.51 |  |
|  | Neighbor | 16.03 | 5.59 |  |
|  | Other | 2.24 | 5.90 |  |

P value of chi square test presented in table 2 also shows that source of knowledge of the people in urban area doesn’t differ significantly with the source of knowledge of the people in rural area in Bangladesh.

Table 3 summarize the participants according to their knowledge on transmission. We have conducted Chi Square test to check the association between the knowledge of urban and rural people in Bangladesh. COVID-19 spreads from person-to-person through several mediums. Knowledge of urban and rural people about these mediums are associated at 5% level of significance except the sneezing and the surface they have recently touched. Their knowledge on food including refrigerated or frozen food, whether COVID-19 spread through food or not, also associated. This result is found to be statistically significant at 1% level of significance. The opinion of warm weather can stop the outbreak of spreading of this virus is now talk of the town. We have found a significant association between the knowledge of urban and rural people about this opinion. Some people are agreed about the opinion of transmission from pet or another animal, where some others are disagreed. Urban and rural people’s knowledge about this transmission is found to be significant. Most of the people in urban area believes that someone can spread the virus without being sick, where most of the rural people bear reverse belief on their mind. This result is also found to be statistically significant at 1% level of significance.

**Table 3:** Population according to their knowledge on TRANSMISSION

| Knowledge of transmission | Urban (n=322) |  | Rural (n=312) |  | P value |
| --- | --- | --- | --- | --- | --- |
|  | Number | % | Number | % |  |
| Spread from person-to-person |  |  |  |  |  |

<sup>1</sup> Heard about corona before attending the survey?
<sup>2</sup> What is the novel corona (COVID-19)?
<sup>3</sup> What are the causes of COVID-19?

|  |  |  |  |  |  |
| --- | --- | --- | --- | --- | --- |
| Coughing | 256 | 79.5 | 220 | 70.5 | 0.012 |
| Sneezing | 233 | 72.4 | 205 | 65.7 | 0.084 |
| Direct contact with a sick person | 249 | 77.3 | 193 | 61.9 | 0.000 |
| The surfaces they have recently touched | 177 | 55.0 | 153 | 49.0 | 0.157 |
| Not known | 15 | 4.7 | 5 | 1.6 | 0.048 |
| Spread through food including refrigerated or frozen food |  |  |  |  |  |
| Yes | 82 | 25.5 | 110 | 35.3 | 0.000 |
| No | 132 | 41.0 | 90 | 28.8 |  |
| There is no evidence support | 108 | 33.5 | 112 | 35.9 |  |
| Warm weather can stop the outbreak |  |  |  |  |  |
| Yes | 63 | 19.6 | 127 | 40.7 | 0.000 |
| No | 120 | 37.3 | 80 | 25.6 |  |
| Yet not confirm | 139 | 43.2 | 105 | 33.7 |  |
| Survive in the surface of the opened things |  |  |  |  |  |
| Few hours | 167 | 51.9 | 88 | 28.2 | 0.000 |
| Few days | 75 | 23.3 | 97 | 31.1 |  |
| Not known | 80 | 24.8 | 127 | 40.7 |  |
| Transmission from pet or another animal |  |  |  |  |  |
| Agree | 221 | 68.6 | 152 | 48.7 | 0.000 |
| Disagree | 101 | 31.4 | 160 | 51.3 |  |
| Someone spread the virus without being sick |  |  |  |  |  |
| Yes | 220 | 68.3 | 122 | 39.1 | 0.000 |
| No | 102 | 31.7 | 190 | 60.9 |  |

Table 4 summarize participants according to their knowledge on sign & symptoms. We have found significant association between the knowledge of urban and rural people on sign & symptoms of COVID-19 except few scenarios. The knowledge about the symptoms that they are generally looking like pneumonia insignificantly varies from urban to rural people. We haven’t found any significant association between the knowledge of urban and rural people on Adults and Older Persons, that they are at higher risk of COVID-19. A small portion of people in both areas, more specifically 0.93% and 0.32%, believes that they could mixed with mass people even if they are the positive cases of COVID-19.

**Table 4:** Population according to their knowledge on SIGN & SYMPTOMS

| Knowledge of SIGN & SYMPTOMS | Urban (n=322) |  | Rural (n=312) |  | P value |
| --- | --- | --- | --- | --- | --- |
|  | Number | % | Number | % |  |
| Incubation period of corona virus is – |  |  |  |  |  |
| 1-2 days | 8 | 2.5 | 5 | 1.6 | 0.000 |
| 2-14 days | 293 | 91.0 | 243 | 77.9 |  |
| One month | 1 | 0.3 | 4 | 1.3 |  |
| Not known | 20 | 6.2 | 60 | 19.2 |  |
| The general symptoms are |  |  |  |  |  |
| Sore throat | 243 | 75.5 | 212 | 67.9 | 0.044 |
| Runny nose | 212 | 65.8 | 170 | 54.5 | 0.004 |
| Nasal congestion | 170 | 52.8 | 117 | 37.5 | 0.000 |
| Shortness of breath | 251 | 78.0 | 175 | 56.1 | 0.000 |
| Difficulty breathing | 252 | 78.3 | 201 | 64.4 | 0.000 |
| Fatigue | 125 | 38.8 | 88 | 28.2 | 0.006 |
| Diarrhea | 107 | 33.2 | 141 | 45.2 | 0.002 |
| The symptoms are generally looking like pneumonia |  |  |  |  |  |
| Yes | 298 | 92.5 | 280 | 89.7 | 0.269 |
| No | 24 | 7.5 | 32 | 10.3 |  |
| Higher risk for COVID-19 |  |  |  |  |  |
| Children | 35 | 10.9 | 90 | 28.8 | 0.000 |
| Adult | 29 | 9.0 | 40 | 12.8 | 0.157 |
| Older persons | 111 | 34.5 | 108 | 34.6 | 0.990 |
| People aged 65 years and older | 189 | 58.7 | 130 | 41.7 | 0.000 |
| Who have serious underlying medical conditions | 218 | 67.7 | 104 | 33.3 | 0.000 |
| I have symptoms of COVID-19 |  |  |  |  |  |
| Self-isolate | 301 | 93.5 | 274 | 87.8 | 0.020 |
| Do not visit a hospital, physician's office,<br>lab or healthcare facility without consulting | 124 | 38.5 | 98 | 31.4 | 0.073 |
| Mixed with mass people | 3 | 0.9 | 1 | 0.3 | 0.638 |

Table 5 summarize the participants according to their knowledge on precautions. The knowledge of urban and rural people on precautions of COVID-19 is found to be significant in most of the cases. Knowledge on the differences between ‘isolation and quarantine’ are not associated in urban and rural area, where majority of them are known with this. All the people in both areas are agreed that if someone recently returned home from abroad should be self-isolate for 14 days after the date of return and monitor for symptoms.

**Table 5:**
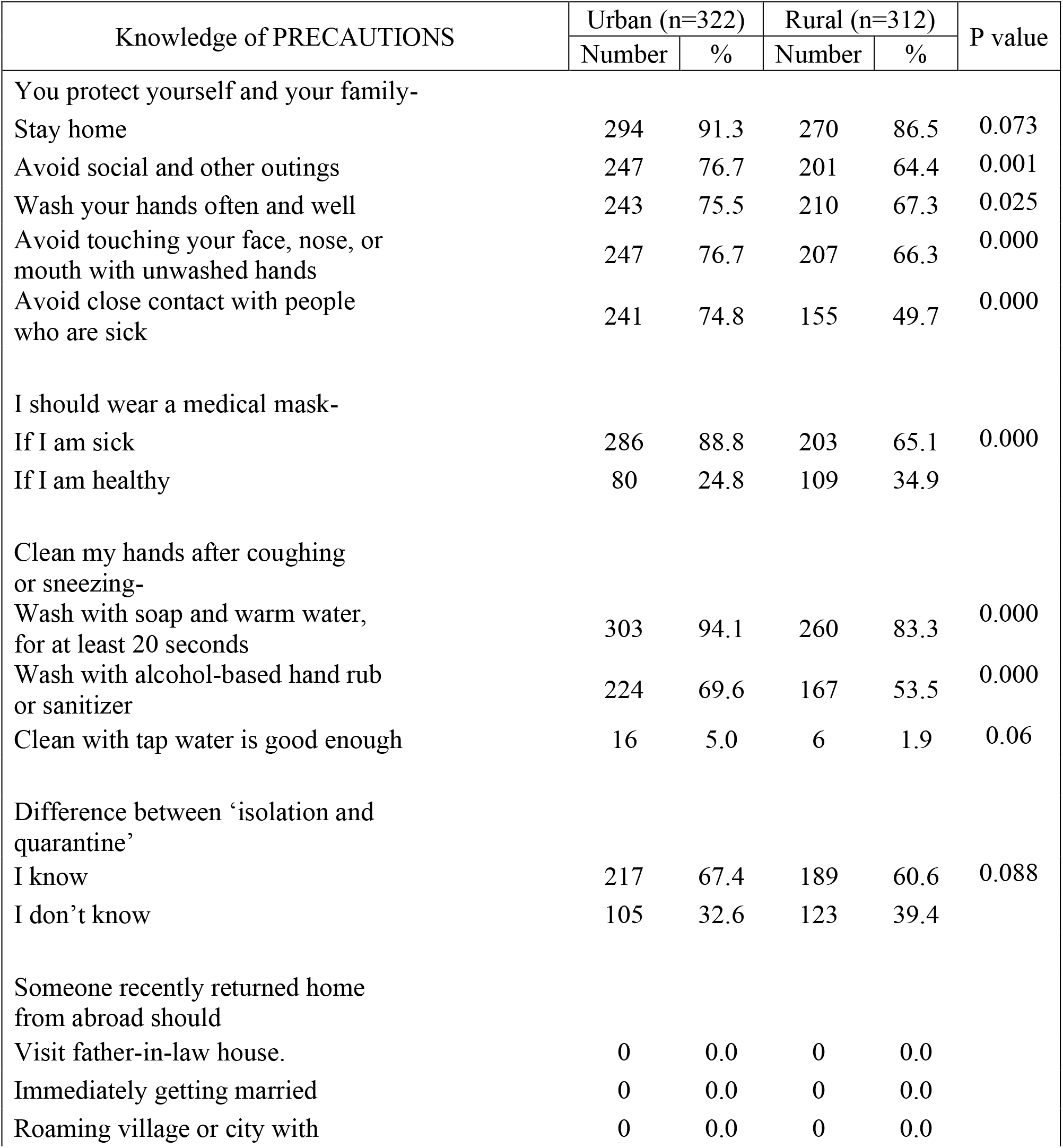
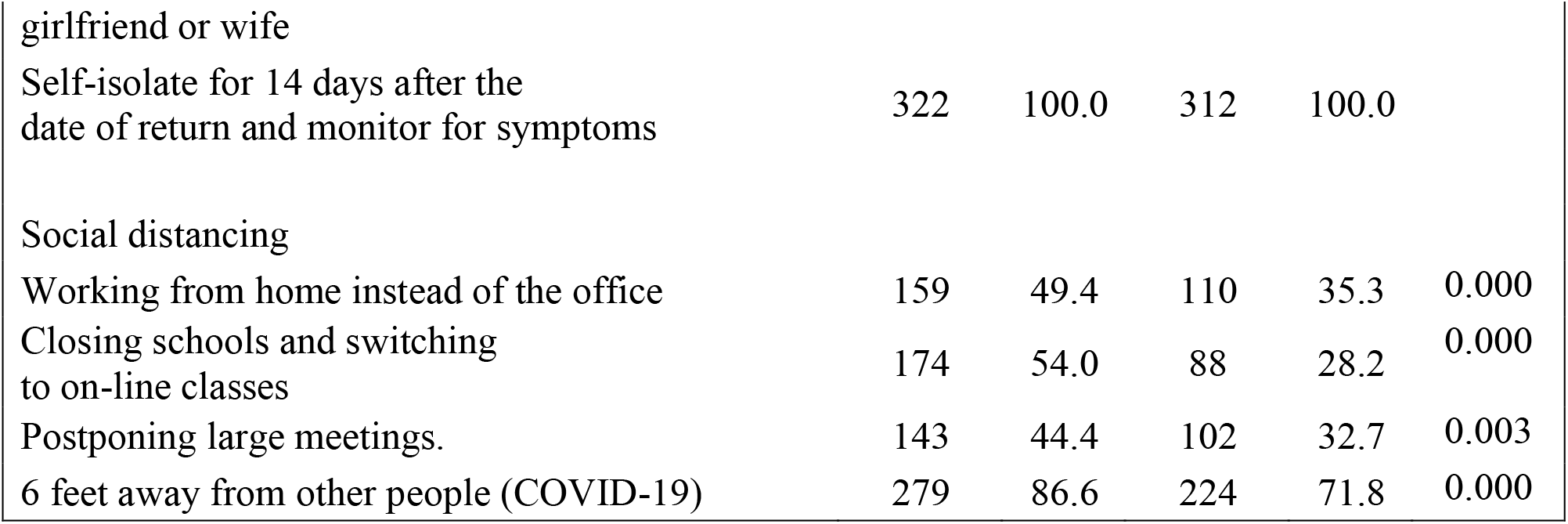
Population according to their knowledge on PRECAUTIONS

Table 6 summarize the participants according to their knowledge on treatment. A large portion of the people in both areas know that there is no suitable treatment or vaccine or anti-biotic for COVID-19. Their knowledge on suitable treatment is significantly associated, where knowledge on vaccine and anti-biotic aren’t. Patients of several diseases are much more vulnerable from this virus. Knowledge about hypertension, asthma and COPD, psychiatric, cardiac, kidney failure, and other patients except diabetic patients are significantly associated in both areas.

**Table 6:**
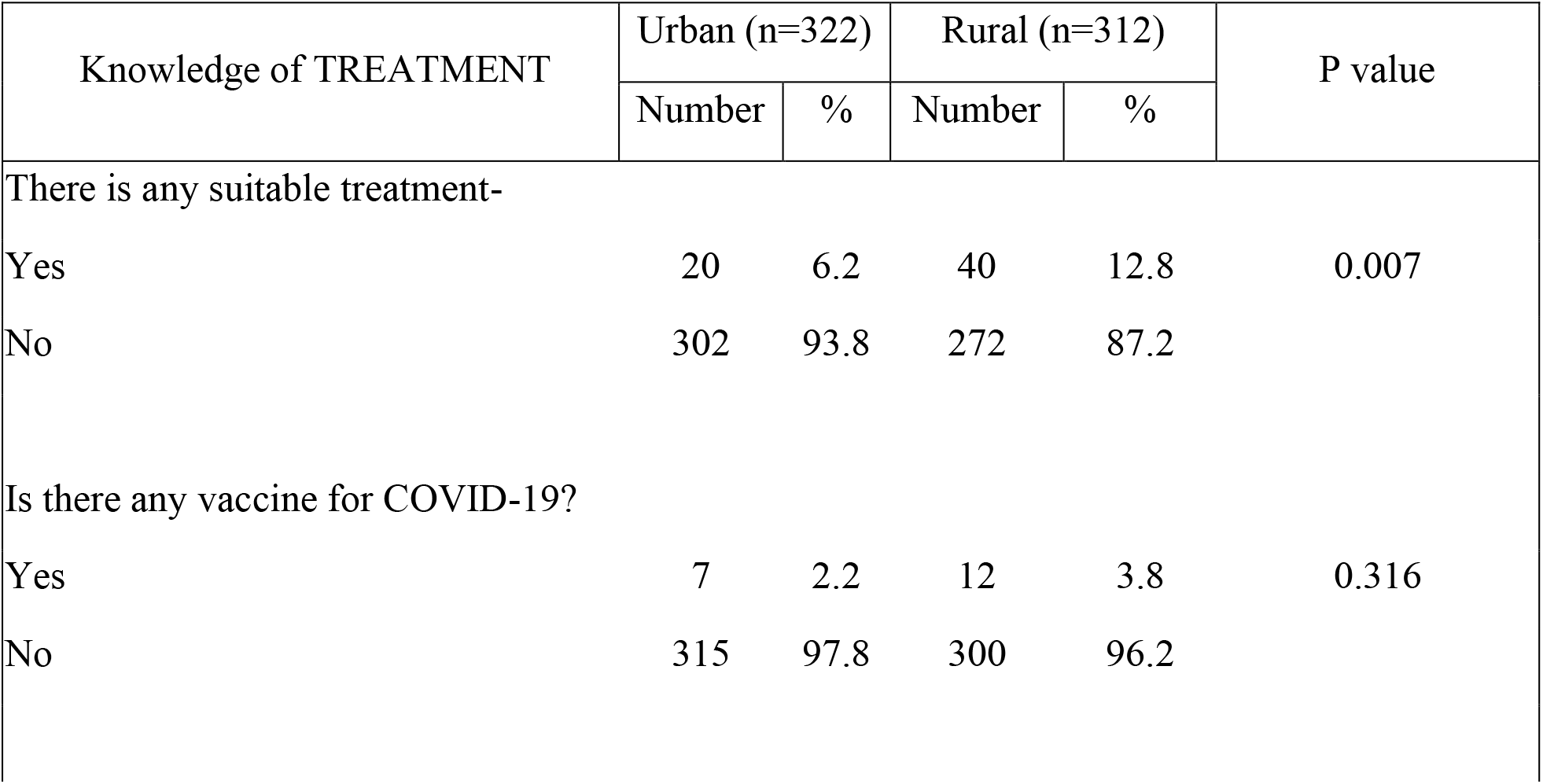
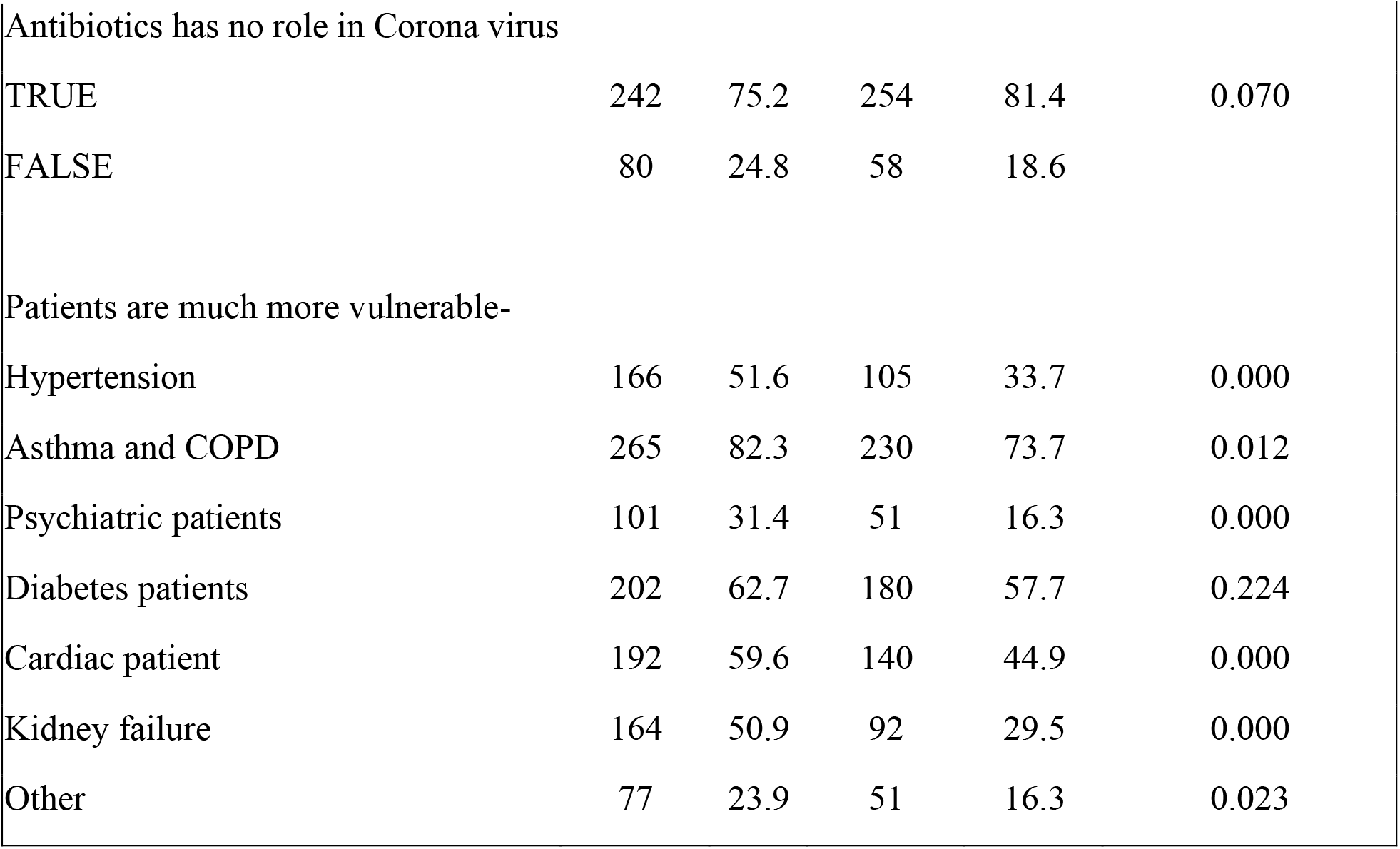
Population according to their knowledge on TREATMENT

| Knowledge of TREATMENT | Urban (n=322) |  | Rural (n=312) |  | P value |
| --- | --- | --- | --- | --- | --- |
|  | Number | % | Number | % |  |
| There is any suitable treatment- |  |  |  |  |  |
| Yes | 20 | 6.2 | 40 | 12.8 | 0.007 |
| No | 302 | 93.8 | 272 | 87.2 |  |
| Is there any vaccine for COVID-19? |  |  |  |  |  |
| Yes | 7 | 2.2 | 12 | 3.8 | 0.316 |
| No | 315 | 97.8 | 300 | 96.2 |  |
| Antibiotics has no role in Corona virus |  |  |  |  |  |
| TRUE | 242 | 75.2 | 254 | 81.4 | 0.070 |
| FALSE | 80 | 24.8 | 58 | 18.6 |  |
| Patients are much more vulnerable- |  |  |  |  |  |
| Hypertension | 166 | 51.6 | 105 | 33.7 | 0.000 |
| Asthma and COPD | 265 | 82.3 | 230 | 73.7 | 0.012 |
| Psychiatric patients | 101 | 31.4 | 51 | 16.3 | 0.000 |
| Diabetes patients | 202 | 62.7 | 180 | 57.7 | 0.224 |
| Cardiac patient | 192 | 59.6 | 140 | 44.9 | 0.000 |
| Kidney failure | 164 | 50.9 | 92 | 29.5 | 0.000 |
| Other | 77 | 23.9 | 51 | 16.3 | 0.023 |

Table 7 summarize the participants according to effect on mental health. All the questions in this section are Likert questions except the last one. We have conducted the Cochran–Armitage test to check the association. This association test can be performed on a contingency table with one ordered nominal variable and one non-ordered nominal variable. The effect of COVID-19 on the mental health of urban and rural people in Bangladesh is found to be significantly associated from many dimensions.

**Table 7:** Population according to effect on MENTAL HEALTH

| Effects on MENTAL HEALTH | Urban (n=322) |  | Rural (n=312) |  | P value |
| --- | --- | --- | --- | --- | --- |
|  | Number | % | Number | % |  |
| I am likely to get Coronavirus |  |  |  |  |  |
| Strongly disagree | 117 | 36.3 | 106 | 34.0 | 0.003 |
| Disagree | 119 | 37.0 | 78 | 25.0 |  |
| Agree | 71 | 22.0 | 116 | 37.2 |  |
| Strongly agree | 15 | 4.7 | 12 | 3.8 |  |
| Worried about getting the Coronavirus |  |  |  |  |  |
| Strongly disagree | 29 | 9.0 | 21 | 6.7 | 0.010 |
| Disagree | 43 | 13.4 | 33 | 10.6 |  |
| Agree | 160 | 49.7 | 131 | 42.0 |  |
| Strongly agree | 90 | 28.0 | 127 | 40.7 |  |
| Belief regarding the infected people- |  |  |  |  |  |
| Will die | 32 | 9.9 | 29 | 9.3 | 0.333 |
| No comments | 162 | 50.3 | 176 | 56.4 |  |
| Will recover | 128 | 39.8 | 107 | 34.3 |  |
| Religious habits change during outbreak |  |  |  |  |  |
| Increased | 203 | 63.0 | 146 | 46.8 | 0.000 |
| No change | 88 | 27.3 | 131 | 42.0 |  |
| Decreased | 31 | 9.6 | 35 | 11.2 |  |
| Life will be changed in a major way<br>after outbreak |  |  |  |  |  |
| Agree | 285 | 88.5 | 259 | 83.0 | 0.062 |
| Disagree | 37 | 11.5 | 53 | 17.0 |  |
| Everyone should follow the government instructions |  |  |  |  |  |
| Strongly disagree | 2 | 0.6 | 20 | 6.4 | 0.000 |
| Disagree | 3 | 0.9 | 6 | 1.9 |  |
| Agree | 42 | 13.0 | 66 | 21.2 |  |
| Strongly agree | 275 | 85.4 | 220 | 70.5 |  |
| Feeling Comfort going to outside of home |  |  |  |  |  |
| Yes | 20 | 6.2 | 42 | 13.5 | 0.003 |
| No | 302 | 93.8 | 270 | 86.5 |  |

## Discussion

Our study shows that urban people are more knowledgeable than rural people regarding causes of COVID-19. This is because, the people in urban and rural area in Bangladesh doesn’t have the similar access to gather knowledge on various issues. Though mass media is the largest source of knowing about COVID-19 here, rural people are mostly dependent on the broadcast media and their neighbors. However, urban people are found to be more aware about this virus than the rural one. They are mostly dependent on internet, especially social sites, scientific journals, broadcast media etc. These areas are the sources of knowledge about COVID-19 to them. Urban and rural residents of the China, where the first COVID-19 patient was reported, have the moderate level of COVID-19 knowledge and they show a positive attitude toward the disease [30].The urban respondents of Pakistan, a nearby country of Bangladesh, had higher knowledge about COVID-19 disease as compared to rural respondents. Their hygienic behavior was better than rural respondents [31]. However, our data also represents that rural people significantly carry lack of knowledge about transmission as well as precautions. On the other hand, both urban and rural people know that there is no treatment of this disease but rural people are not taking much precautions, our thinking this may be due to the lack knowledge and awareness. Choi and Kim also described in their studies infection-control knowledge directly related with attitudes and practice [21].Ajzen and Fishbein revealed a significant correlation among knowledge, attitudes and practice [28]. Other studies also support this finding where they mentioned lack of awareness in Anhui province of China [29]. Impact of this pandemic on the global health and mental health is reported in recent studies [28]. Our study also shows that both urban and rural people are worried about getting the corona virus and believe that after outbreak life will be changed in a major way. Some researchers tried to identify the root cause of panic in the community, where they reported that the Muslim communities in the rural area facing the COVID-19 Pandemic attempts to find refuge from the plague and hope for survival [32]. However urban people significantly increased their religious habit during this outbreak.

So far, we know, previously population-based studies regarding COVID-19 either field based or online based were conducted in the city area only. In our country, internet is not easily accessible, especially in rural area. Moreover, the people in the rural area are not habituated regarding this on-line survey. Face to face survey is highly recommended here to find out the exact scenario.

In contrast we conducted our study in both urban and rural areas following heath guidelines strictly and also covered a good number of participants in both areas. On the other hand, the limitation of our study, we can’t include survivor in this study because survivor wasn’t available at that time. Our recommendation to conduct similar studies in different developing countries in the world for more evidence for the management of pandemic because rural people is less advanced in different perspectives than urban people.

COVID-19 is now a pandemic and the situation are like time bomb as no medical treatment is too much effective and no proper vaccine yet discovered. Bangladesh become a highest worse condition regarding this infectious disease in the world due to overpopulated and lack of awareness. In this circumstance only people’s awareness in Bangladesh can help to protect us. This cross-sectional study was carried out to identify the awareness and behavioral perspective on COVID-19 between urban and rural people. Awareness regarding COVID-19 was unsatisfactory in rural residents as compared to urban. Based on the results of our study, we can conclude that improvement of COVID-19 knowledge, attitudes and promotion of awareness among residents by effective health education programs is needed especial care needed in rural areas. Our belief this survey will provide valuable information to the legislators regarding the perceptions of urban and rural population for the management of pandemic.

## Data Availability

Data can be shared upon request.

## Acknowledgements

There was no financial and material support in this study.

## Compliance with ethical standards

### Ethical approval

The study was approved by the research ethics committeeof the Department of Pharmacy, Mawlana Bhashani Science and Technology University.

